# Multimodal Sleep Physiology Reconstructs Cerebrospinal Fluid Dynamics: A Candidate Digital Biomarker from Noninvasive Sensing

**DOI:** 10.64898/2026.07.27.26359039

**Authors:** Anton R. Banta, A. Roy Phillips, Raimondo D’Ambrosio, Christof Karmonik, Eugene Golanov, Angelique Regnier-Golanov, Fidaa Shaib, Gavin W. Britz, Behnaam Aazhang

## Abstract

Cerebrospinal fluid (CSF) dynamics are clinically important but can presently be measured only with expensive, non-portable imaging or invasive monitoring, precluding scalable assessment. We investigated whether CSF flow in the cerebral aqueduct can be reconstructed from noninvasive physiology recorded during sleep. In 22 healthy adults, with independent validation in 14 additional adults, regularized linear and ensemble regressors predicted the cardiac-cycle CSF flow waveform from electrocardiography, photoplethysmography (PPG), respiration, and electroencephalography acquired during overnight polysomnography, using 7-Tesla phase-contrast MRI as ground truth. Waveforms were reconstructed with high fidelity and generalized to the validation cohort, and deriving physiological metrics from the reconstructed waveform estimated peak velocity more accurately than predicting it directly. A minimal, wearable-oriented PPG feature set retained performance. These results support the feasibility of a noninvasive, potentially wearable digital biomarker of CSF dynamics and motivate evaluation in neurological disease populations.

## Introduction

Cerebrospinal fluid (CSF) plays a central role in maintaining brain homeostasis, facilitating nutrient transport, metabolic waste clearance, and mechanical protection of neural tissue^1^. Dysregulation of CSF dynamics has been implicated in a range of neurological conditions, including hydrocephalus, Chiari malformation, intracranial hypertension, and neurodegenerative diseases associated with impaired glymphatic clearance^2–6^. Despite its importance, CSF flow remains difficult to measure in a scalable and noninvasive manner in humans.

Current approaches for assessing CSF dynamics rely primarily on phase-contrast magnetic resonance imaging (PC-MRI), which enables quantification of flow velocity and volumetric flux in structures such as the cerebral aqueduct^7,8^. While effective, MRI-based measurements are expensive, non-portable, and provide only sparse snapshots in time, limiting their utility for longitudinal monitoring or large-scale studies. Invasive approaches, such as ventriculostomy, provide direct measurements but are not suitable outside of acute clinical settings^9^. As a result, there is a critical need for scalable methods to assess CSF dynamics noninvasively.

Recent advances in digital medicine have enabled continuous acquisition of physiological signals using noninvasive sensors, including electroencephalography (EEG), electrocardiography (ECG), photoplethysmography (PPG), and respiratory monitoring. These modalities capture neural, cardiovascular, and respiratory processes that are known to influence CSF movement, including arterial pulsatility, respiration-driven pressure changes, and sleep-dependent neural activity^4,10^. Prior studies have demonstrated coupling between electrophysiological activity, vascular dynamics, and CSF flow, particularly during sleep^11,12^. However, it remains unclear whether these signals contain sufficient information to infer CSF dynamics at the individual level.

To date, few studies have attempted to estimate CSF flow without direct imaging, and existing approaches are typically limited to single modalities or indirect proxies of CSF dynamics^13–15^. Moreover, prior work has largely focused on predicting summary metrics rather than reconstructing the underlying temporal dynamics of CSF flow. Whether multimodal physiological signals can be integrated to recover the full CSF waveform, and whether such representations provide advantages over direct metric prediction, remains an open question.

In this study, we investigate whether CSF flow in the cerebral aqueduct can be reconstructed from multimodal physiological measurements acquired during sleep. We collected overnight polysomnography, including EEG, ECG, PPG, respiration, sleep staging, and body position, together with 7T PC-MRI measurements of CSF flow in a cohort of healthy adults, with the two acquisitions performed in separate sessions. Using regularized linear and ensemble regressors (ridge regression and random forest), we trained predictors to estimate both the full CSF flow waveform and derived physiological metrics. Model performance was evaluated using a leave-one-out framework in a development cohort (n=22) and further assessed in an independent validation cohort (n=14).

We demonstrate that CSF waveforms can be reconstructed from noninvasive physiological signals with high fidelity and that deriving physiological metrics from reconstructed waveforms yields more accurate estimates than predicting those metrics directly. These findings suggest that preserving the temporal dynamics of physiological systems may be critical for accurate inference and support the feasibility of a digital biomarker of CSF dynamics. Such an approach, achievable even from a minimal set of features obtainable from consumer wearable sensors, could enable scalable, noninvasive monitoring of brain fluid physiology and motivate future studies in neurological disease populations.

## Results

### Study participants and overview

The development cohort comprised 22 healthy adults (mean age 27 ± 4 years, range 22–39; 12 male, 10 female). An independent validation cohort of 14 healthy adults (mean age 29 ± 5 years, range 21–35; 7 male, 7 female) was recruited at a later period and processed entirely by a third party; this cohort was withheld from all modelling and hyperparameter decisions and used exclusively for external validation. 7T PC-MRI provided 20-phase cardiac-cycle waveforms of cerebrospinal fluid (CSF) flow through the cerebral aqueduct as the ground-truth target. Noninvasive overnight polysomnography (PSG) yielded six feature sets: ECG (110 features), PPG (80 features), oronasal respiration (Resp; 85 features), EEG (2,170 features), a physiologically naïve heart-rate and respiration-rate baseline (naive-HR; 10 features), and a full-modality fusion (ECG + PPG + RES + EEG concatenated; 2,445 features). All features were computed separately for each of five sleep stages (Wake, N1, N2, N3, REM). Participant demographics for both cohorts are summarized in Table 1, and the overall acquisition and modelling workflow is illustrated in Fig. 1.

**Fig. 1.**
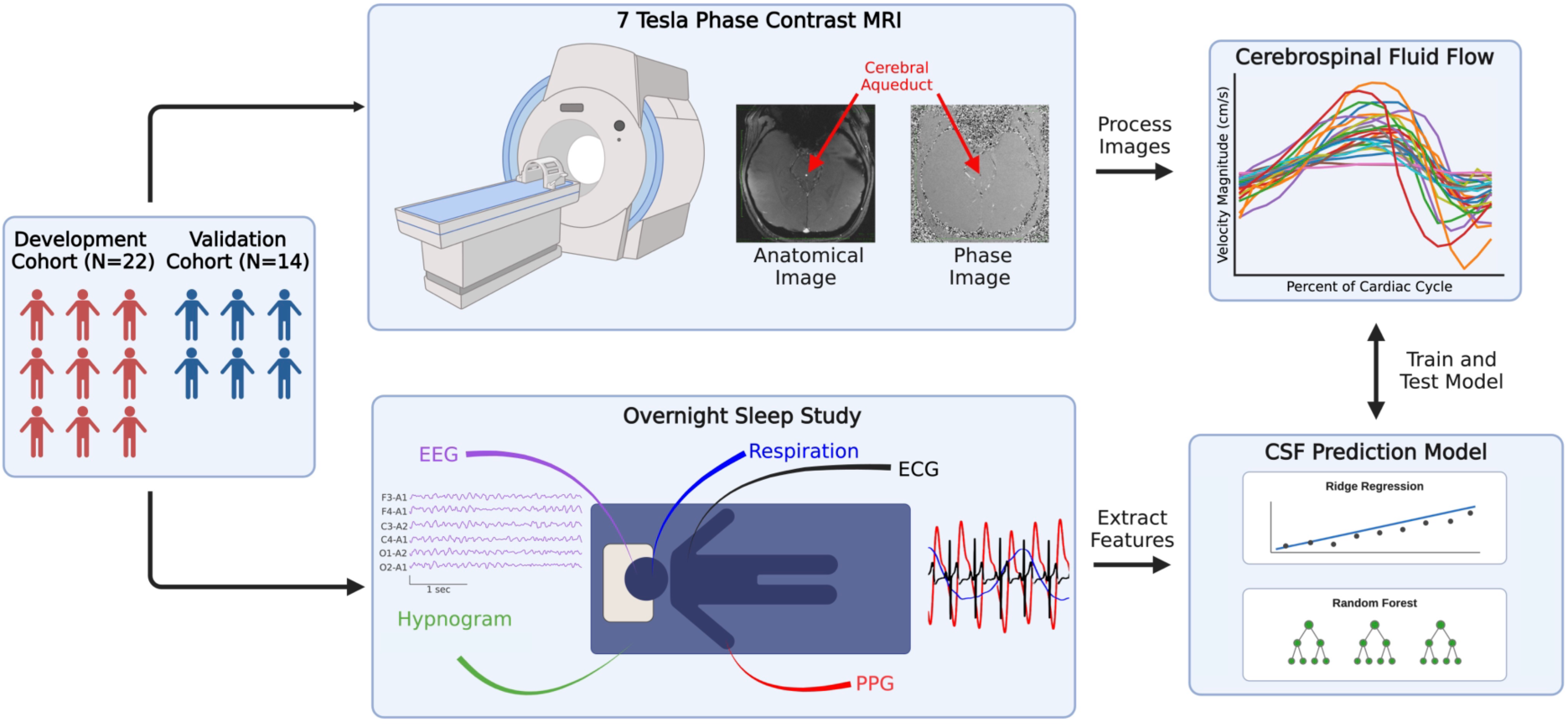
Overview of the study workflow for reconstructing MRI-derived cerebrospinal fluid dynamics from noninvasive sleep physiology. Healthy adult participants were enrolled across two independent cohorts: a development cohort (*N* = 22) used for model training and leave-one-out cross-validation, and a separate validation cohort (*N* = 14) used to assess generalization. Each participant underwent two acquisitions. First, 7-Tesla phase-contrast MRI was used to image the cerebral aqueduct, yielding anatomical and velocity-encoded phase images from which the ground-truth cerebrospinal fluid (CSF) flow and velocity waveform was derived across the cardiac cycle. Second, an overnight polysomnography study captured multimodal physiological signals, including electroencephalography (EEG), electrocardiography (ECG), photoplethysmography (PPG), respiration, and hypnogram-derived sleep stage and body position. Features were extracted from each physiological modality and used as inputs to machine-learning regression models (ridge regression and random forest) trained to predict the CSF flow/velocity waveform. Predicted waveforms were compared against the MRI-derived ground truth to train and evaluate model performance.

**Table 1.** Participant Information.

| Characteristic | Development Cohort | Validation Cohort |
| --- | --- | --- |
| Number of Participants | 22 | 14 |
| Age (years) | 27 ± 4 (22-39) | 29 ± 5 (21-35) |
| Male:Female | 12:10 | 7:7 |
| Age Group 20-29 | 17 | 7 |
| Age Group 30-39 | 5 | 7 |

Demographic characteristics of the development and validation cohorts, including the number of participants, age (mean ± SD with range), sex distribution, and age-group breakdown.

### CSF waveform reconstruction from noninvasive sleep physiology

Both regularized linear (ridge) and ensemble (random forest, RF) regressors reconstructed the 20-point cardiac-cycle CSF flow waveform from each noninvasive feature set in the development cohort (Table 2). Waveform agreement was uniformly high and tightly clustered across modalities: for ridge regression, the median Pearson correlation between predicted and ground-truth waveforms ranged from 0.94 to 0.96 across the six feature sets, with the multimodal fusion model reaching a median r of 0.95 (IQR 0.93–0.97). Representative reconstructions spanning the worst-, median-, and best-performing subjects are shown in Fig. 2; in the median and best cases both models recovered the overall morphology and timing of the waveform, whereas in the worst-performing subjects the predicted waveform preserved timing but systematically underestimated peak amplitude.

**Fig. 2.**
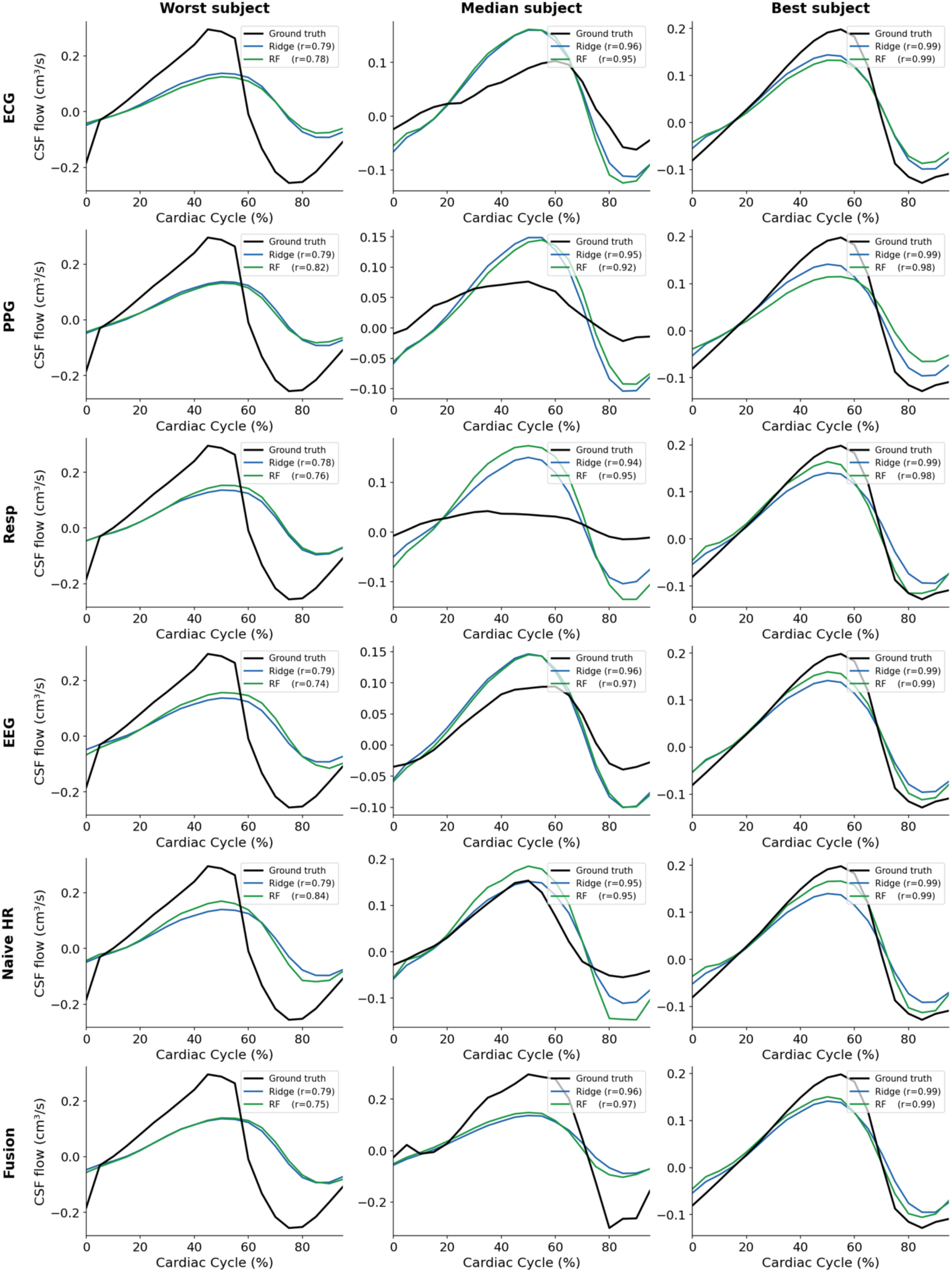
Representative reconstructions of the cerebrospinal fluid flow waveform across physiological modalities and models for the development cohort. Each panel shows the MRI-derived ground-truth cerebrospinal fluid (CSF) flow waveform (black) over the cardiac cycle alongside reconstructions from ridge regression (blue) and random forest (green), with the per-panel Pearson correlation coefficient (*r*) between each predicted and ground-truth waveform reported in the legend. Rows correspond to the input modality used for prediction: electrocardiography (ECG), photoplethysmography (PPG), respiration (Resp), electroencephalography (EEG), a naïve heart-rate baseline (Naïve HR), and the multimodal fusion model combining all modalities (Fusion). Columns show representative participants ranked by reconstruction accuracy (Pearson *r*) — the worst-, median-, and best-performing subject — to illustrate the range of model behavior. CSF flow is expressed in cm³/s and the cardiac cycle as a percentage from 0 to 100%. Across modalities, both models recover the overall morphology and timing of the CSF waveform in the median-and best-performing subjects, whereas reconstructions in the worst-performing subjects preserve waveform timing but systematically underestimate peak amplitude.

**Table 2.** Waveform Agreement for Development and Validation Cohorts.

| **Model** | **Modality** | **Dev Pearson r** | **Val Pearson r** | **Dev CCC** | **Val CCC** | **Dev nRMSE** | **Val nRMSE** |
| --- | --- | --- | --- | --- | --- | --- | --- |
| **Ridge** | ECG | 0.96 (0.93–0.98) | 0.97 (0.87–0.99) | 0.81 (0.71–0.93) | 0.86 (0.61–0.94) | 0.20 (0.14–0.29) | 0.17 (0.13–0.28) |
| **Ridge** | PPG | 0.95 (0.92–0.97) | 0.96 (0.83–0.98) | 0.82 (0.68–0.93) | 0.87 (0.60–0.94) | 0.19 (0.14–0.28) | 0.16 (0.12–0.29) |
| **Ridge** | Resp | 0.94 (0.92–0.96) | 0.95 (0.83–0.98) | 0.80 (0.69–0.92) | 0.88 (0.59–0.95) | 0.20 (0.16–0.28) | 0.17 (0.13–0.29) |
| **Ridge** | EEG | 0.95 (0.92–0.97) | 0.96 (0.83–0.98) | 0.83 (0.68–0.93) | 0.87 (0.59–0.94) | 0.19 (0.14–0.28) | 0.17 (0.12–0.29) |
| **Ridge** | Naive HR | 0.95 (0.91–0.97) | 0.95 (0.82–0.98) | 0.80 (0.68–0.92) | 0.89 (0.57–0.93) | 0.21 (0.15–0.27) | 0.17 (0.13–0.29) |
| **Ridge** | Fusion | 0.95 (0.93–0.97) | 0.96 (0.84–0.98) | 0.81 (0.69–0.93) | 0.87 (0.59–0.95) | 0.19 (0.14–0.28) | 0.17 (0.13–0.29) |
| **RF** | ECG | 0.95 (0.91–0.98) | 0.98 (0.88–0.98) | 0.74 (0.65–0.90) | 0.90 (0.62–0.96) | 0.21 (0.18–0.41) | 0.17 (0.10–0.27) |
| **RF** | PPG | 0.95 (0.92–0.98) | 0.93 (0.89–0.97) | 0.84 (0.71–0.91) | 0.80 (0.71–0.93) | 0.20 (0.15–0.26) | 0.20 (0.14–0.44) |
| **RF** | Resp | 0.95 (0.91–0.97) | 0.95 (0.83–0.98) | 0.82 (0.68–0.90) | 0.90 (0.62–0.94) | 0.19 (0.16–0.26) | 0.17 (0.12–0.27) |
| **RF** | EEG | 0.96 (0.89–0.98) | 0.94 (0.83–0.97) | 0.80 (0.65–0.92) | 0.85 (0.59–0.86) | 0.19 (0.14–0.30) | 0.22 (0.18–0.32) |
| **RF** | Naive HR | 0.96 (0.94–0.98) | 0.96 (0.87–0.99) | 0.82 (0.70–0.89) | 0.88 (0.60–0.92) | 0.21 (0.18–0.27) | 0.18 (0.12–0.27) |
| **RF** | Fusion | 0.96 (0.91–0.97) | 0.94 (0.87–0.98) | 0.81 (0.68–0.91) | 0.87 (0.60–0.89) | 0.20 (0.17–0.26) | 0.20 (0.16–0.30) |

Per-subject agreement between predicted and MRI-derived ground-truth CSF flow waveforms for ridge regression and random forest (RF) across six input feature sets (ECG, PPG, respiration [Resp], EEG, a naïve heart-rate baseline [Naïve HR], and the multimodal fusion model [Fusion]), quantified by Pearson correlation coefficient (r), Lin’s concordance correlation coefficient (CCC), and range-normalized root-mean-square error (nRMSE). Values are median (interquartile range) across subjects for the development cohort (leave-one-out cross-validation, n = 22) and the independent validation cohort (n = 14).

Because the Pearson correlation is insensitive to differences in amplitude and offset, we additionally report Lin’s concordance correlation coefficient (CCC) and range-normalized root-mean-square error (nRMSE), which penalize amplitude and scale error (Table 2). Median CCC was consistently lower than the corresponding Pearson r (for example, 0.81 versus 0.95 for the ridge fusion model), and median nRMSE was approximately 0.19–0.21, indicating that a residual fraction of subjects exhibited amplitude discrepancies not captured by correlation alone, consistent with the amplitude underestimation visible in the worst-performing subjects of Fig. 2.

Agreement for the derived mean flow was further examined using Bland–Altman analysis (Fig. 3a). Both models showed negligible bias (mean difference near zero across all modalities), with the large majority of subjects falling within the ±1.96 SD limits of agreement. Ridge predictions followed a tight, near-linear negative trend, reflecting the controlled shrinkage of regularized predictions toward the cohort mean, whereas random forest predictions were more dispersed while remaining within the limits of agreement. Equivalent analyses for volume and peak velocity are provided in Supplementary Fig. 1.

**Fig. 3.**
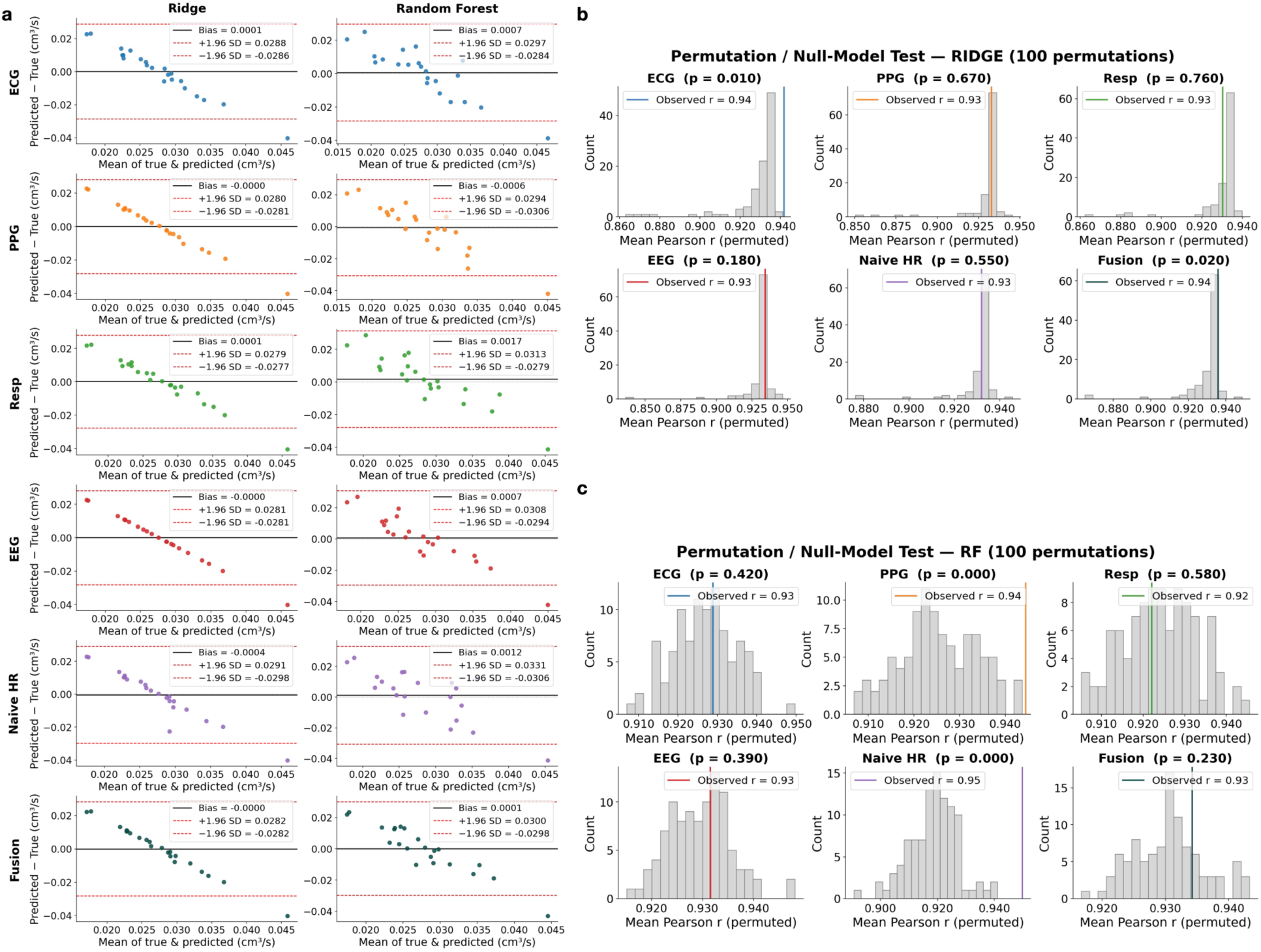
Agreement analysis and permutation-based significance testing of CSF flow reconstructions. **a** Bland–Altman plots comparing waveform-derived mean CSF flow predictions against MRI-derived ground truth for ridge regression (left column) and random forest (right column) across each input modality (rows: ECG, PPG, respiration [Resp], EEG, a naïve heart-rate baseline [Naïve HR], and the multimodal fusion model [Fusion]). In each plot, the difference between predicted and true flow is plotted against their mean; the solid black line denotes the mean bias and the dashed red lines denote the ±1.96 SD limits of agreement. Both models showed negligible bias (near zero across all modalities), with the large majority of points falling within the limits of agreement. Ridge predictions followed a tighter, near-linear negative trend, reflecting controlled shrinkage of predictions toward the cohort mean, whereas random forest predictions showed greater dispersion while remaining within the limits of agreement. **b, c** Permutation/null-model tests for ridge (**b**) and random forest (**c**), each based on 100 permutations in which subject labels were randomly shuffled prior to retraining. For each modality, the grey histogram shows the null distribution of the mean Pearson correlation coefficient (*r*) obtained under permuted labels, the vertical-colored line indicates the observed (unpermuted) mean *r*, and the empirical *p*-value is reported in each subpanel title. Several modalities reached statistical significance — ECG (*p* = 0.010) and fusion (*p* = 0.020) for ridge, and PPG (*p* < 0.001) and the naïve heart-rate baseline (*p* < 0.001) for random forest — despite the modest development-cohort size (*N* = 22), indicating that the models capture subject-specific structure beyond chance.

To confirm that reconstruction accuracy reflected subject-specific information rather than the stereotyped morphology shared across healthy subjects, we performed permutation testing in which subject labels were randomly shuffled prior to retraining (100 permutations; Fig. 3b,c). The null distributions of mean Pearson r were centered at high values (approximately 0.92–0.93), confirming that most of the absolute correlation reflects the common systolic-peak and diastolic-trough shape of the CSF waveform rather than individualized prediction. Nonetheless, several observed values exceeded the null distribution: ECG (p = 0.010) and fusion (p = 0.020) reached significance for ridge regression, and PPG (p < 0.001) and the naïve heart-rate baseline (p < 0.001) for random forest, indicating a statistically significant subject-specific component despite the modest cohort size (n = 22). Notably, the naïve heart-rate baseline performed comparably to the full fusion model on these aggregate waveform metrics, indicating that median waveform agreement is largely saturated by shared cardiac-driven morphology and that modality-specific information is better resolved in the derived metrics examined next. The naïve heart-rate baseline was included throughout as a deliberately minimal control: because cardiac pulsatility is itself a driver of CSF motion, it represents the performance attainable from heart rate alone and provides a reference against which the contribution of richer feature sets can be judged.

### Waveform-first prediction outperforms direct metric estimation for peak velocity

We next asked whether physiologically meaningful CSF metrics (mean flow, volume, and peak (maximum) velocity) are better estimated by deriving them from the reconstructed waveform (waveform-first, Mode 1) or by training models to predict each scalar metric directly (Mode 2). Absolute errors for the waveform-first approach are reported in Table 3; for example, the ridge fusion model achieved a median peak-velocity MAPE of 41.7% in the development cohort. We then compared the two strategies subject-by-subject using the paired difference in error, ΔMAPE = MAPE(Mode 1) − MAPE(Mode 2), with 95% bootstrap confidence intervals (Fig. 4).

**Fig. 4.**
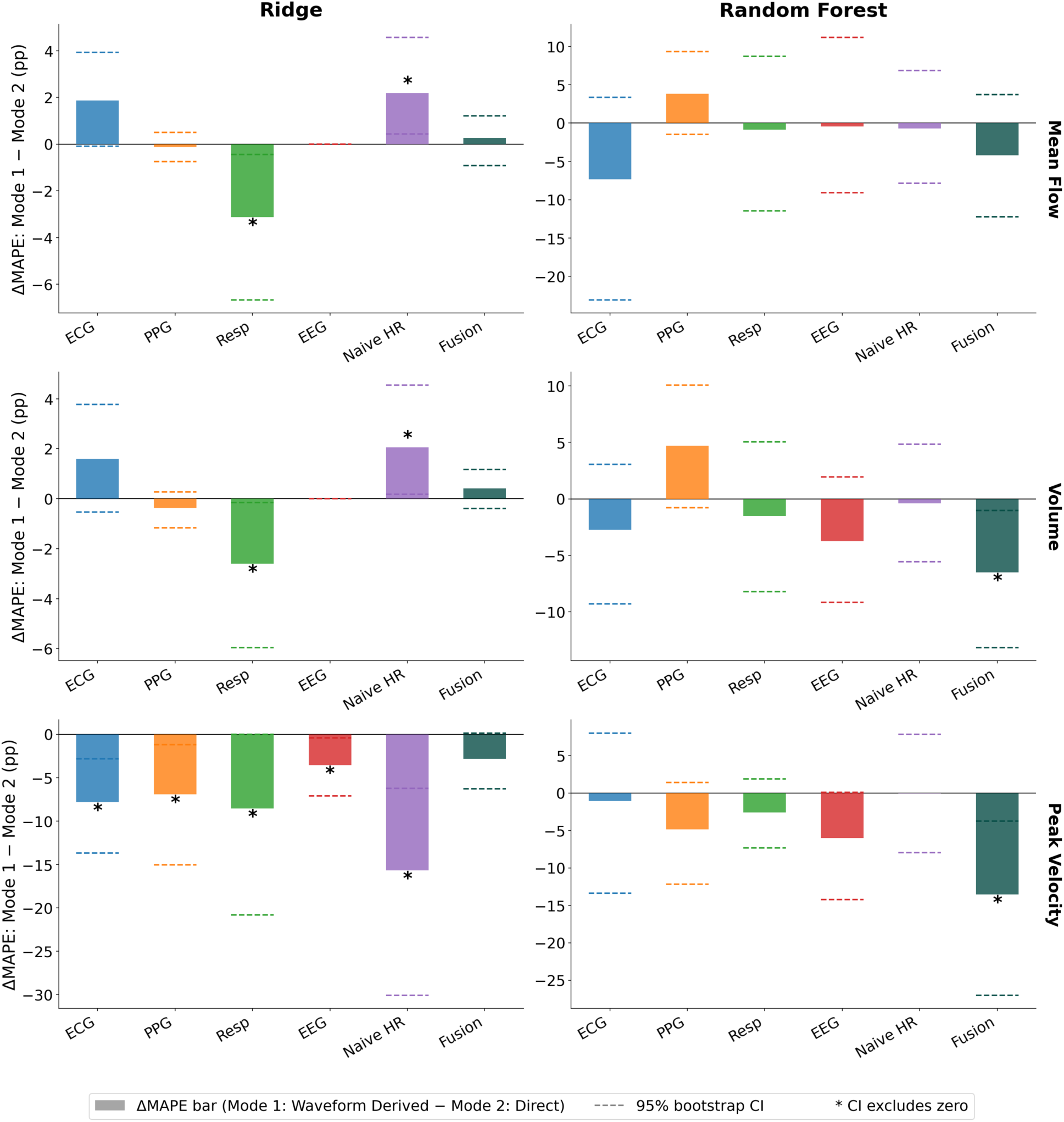
Comparison of waveform-first versus direct metric prediction for CSF-derived physiological metrics. Two strategies for estimating CSF flow metrics were compared: predicting the full CSF flow waveform and then deriving the metric from the reconstructed waveform (Mode 1, waveform-first), versus training a model to predict the metric directly as a scalar (Mode 2, direct prediction). Each bar shows the difference in mean absolute percentage error between the two strategies, ΔMAPE = MAPE(Mode 1) − MAPE(Mode 2), in percentage points (pp), such that **negative values indicate lower error for the waveform-first approach**. Columns correspond to the prediction model (ridge regression, left; random forest, right) and rows to the derived metric (mean flow, top; volume, middle; peak velocity, bottom). Within each subpanel, bars denote the point estimate of ΔMAPE for each input modality (ECG, PPG, respiration [Resp], EEG, the naïve heart-rate baseline [Naïve HR], and the multimodal fusion model [Fusion]); dashed lines indicate the 95% bootstrap confidence interval, and an asterisk marks modalities whose interval excludes zero. For peak velocity under ridge regression, the waveform-first approach reduced error across all individual modalities, reaching significance for ECG, PPG, respiration, EEG, and the naïve heart-rate baseline, whereas differences for mean flow and volume were smaller, inconsistent in direction across modalities, and largely non-significant. Random forest comparisons showed wider confidence intervals and fewer significant differences overall.

**Table 3.**
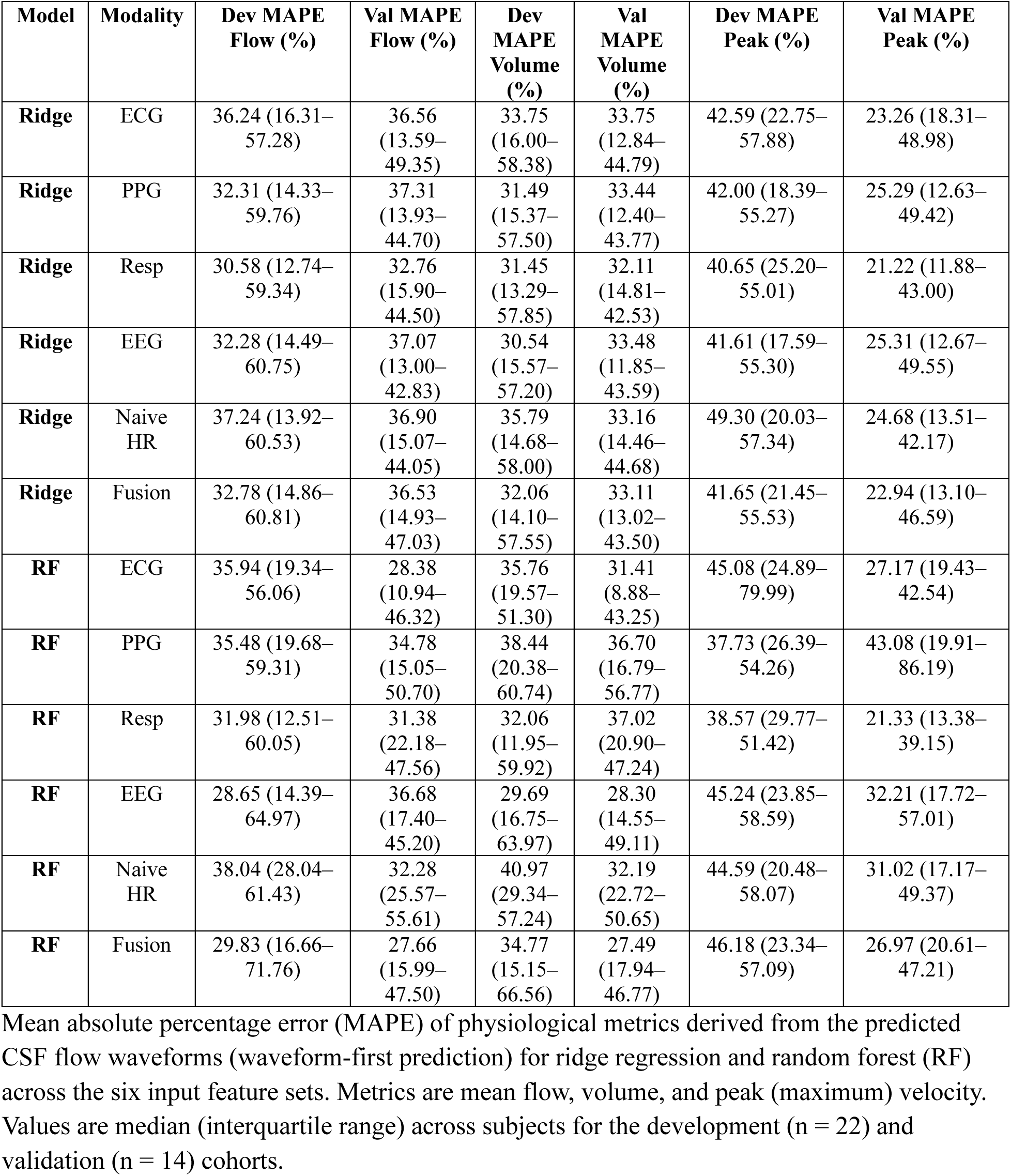
Waveform Metric Agreement for Development and Validation Cohorts.

| **Model** | **Modality** | **Dev MAPE**<br>**Flow (%)** | **Val MAPE**<br>**Flow (%)** | **Dev**<br>**MAPE**<br>**Volume**<br>**(%)** | **Val**<br>**MAPE**<br>**Volume**<br>**(%)** | **Dev MAPE**<br>**Peak (%)** | **Val MAPE**<br>**Peak (%)** |
| --- | --- | --- | --- | --- | --- | --- | --- |
| **Ridge** | ECG | 36.24 (16.31–57.28) | 36.56 (13.59–49.35) | 33.75 (16.00–58.38) | 33.75 (12.84–44.79) | 42.59 (22.75–57.88) | 23.26 (18.31–48.98) |
| **Ridge** | PPG | 32.31 (14.33–59.76) | 37.31 (13.93–44.70) | 31.49 (15.37–57.50) | 33.44 (12.40–43.77) | 42.00 (18.39–55.27) | 25.29 (12.63–49.42) |
| **Ridge** | Resp | 30.58 (12.74–59.34) | 32.76 (15.90–44.50) | 31.45 (13.29–57.85) | 32.11 (14.81–42.53) | 40.65 (25.20–55.01) | 21.22 (11.88–43.00) |
| **Ridge** | EEG | 32.28 (14.49–60.75) | 37.07 (13.00–42.83) | 30.54 (15.57–57.20) | 33.48 (11.85–43.59) | 41.61 (17.59–55.30) | 25.31 (12.67–49.55) |
| **Ridge** | Naive HR | 37.24 (13.92–60.53) | 36.90 (15.07–44.05) | 35.79 (14.68–58.00) | 33.16 (14.46–44.68) | 49.30 (20.03–57.34) | 24.68 (13.51–42.17) |
| **Ridge** | Fusion | 32.78 (14.86–60.81) | 36.53 (14.93–47.03) | 32.06 (14.10–57.55) | 33.11 (13.02–43.50) | 41.65 (21.45–55.53) | 22.94 (13.10–46.59) |
| **RF** | ECG | 35.94 (19.34–56.06) | 28.38 (10.94–46.32) | 35.76 (19.57–51.30) | 31.41 (8.88–43.25) | 45.08 (24.89–79.99) | 27.17 (19.43–42.54) |
| **RF** | PPG | 35.48 (19.68–59.31) | 34.78 (15.05–50.70) | 38.44 (20.38–60.74) | 36.70 (16.79–56.77) | 37.73 (26.39–54.26) | 43.08 (19.91–86.19) |
| **RF** | Resp | 31.98 (12.51–60.05) | 31.38 (22.18–47.56) | 32.06 (11.95–59.92) | 37.02 (20.90–47.24) | 38.57 (29.77–51.42) | 21.33 (13.38–39.15) |
| **RF** | EEG | 28.65 (14.39–64.97) | 36.68 (17.40–45.20) | 29.69 (16.75–63.97) | 28.30 (14.55–49.11) | 45.24 (23.85–58.59) | 32.21 (17.72–57.01) |
| **RF** | Naive HR | 38.04 (28.04–61.43) | 32.28 (25.57–55.61) | 40.97 (29.34–57.24) | 32.19 (22.72–50.65) | 44.59 (20.48–58.07) | 31.02 (17.17–49.37) |
| **RF** | Fusion | 29.83 (16.66–71.76) | 27.66 (15.99–47.50) | 34.77 (15.15–66.56) | 27.49 (17.94–46.77) | 46.18 (23.34–57.09) | 26.97 (20.61–47.21) |

Mean absolute percentage error (MAPE) of physiological metrics derived from the predicted CSF flow waveforms (waveform-first prediction) for ridge regression and random forest (RF) across the six input feature sets. Metrics are mean flow, volume, and peak (maximum) velocity. Values are median (interquartile range) across subjects for the development (n = 22) and validation (n = 14) cohorts.

For peak velocity under ridge regression, the waveform-first approach reduced error for every individual modality, with the confidence interval excluding zero for ECG, PPG, respiration, EEG, and the naïve heart-rate baseline (Fig. 4, bottom-left). In contrast, differences for mean flow and volume were smaller, inconsistent in direction across modalities, and largely non-significant. Random forest comparisons showed the same qualitative pattern but with wider confidence intervals and fewer significant differences.

This dissociation has a straightforward interpretation: peak velocity is a single point on the waveform whose accurate recovery requires the underlying morphology to be correct, and is therefore aided by first reconstructing the full curve. Mean flow and volume, by contrast, are aggregate quantities (an average and an integral over the cardiac cycle) that a direct scalar regressor can approximate without preserving waveform shape. The advantage of waveform-first prediction is thus specific to shape-dependent metrics rather than a general improvement and is most consistent for peak velocity under ridge regression.

### Generalization to an independent validation cohort

To assess generalization, models trained on the development cohort were applied without modification to the independent validation cohort (n = 14), which was processed by a third party and withheld from all modelling decisions. Median waveform agreement was preserved: validation median Pearson r ranged from 0.93 to 0.98, CCC from 0.80 to 0.90, and nRMSE from 0.16 to 0.22 across models and modalities (Table 2). Development and validation distributions were compared for each model and modality using two-sided Mann–Whitney U tests, and no comparison reached significance (Fig. 5), consistent with preserved generalization; given the cohort sizes, this absence of a detected difference should be interpreted as compatible with, rather than proof of, equivalence. The same preservation held for the waveform-derived metrics (mean flow, volume, and peak velocity), which likewise showed no significant development-versus-validation differences (Fig. 5b).

**Fig. 5.**
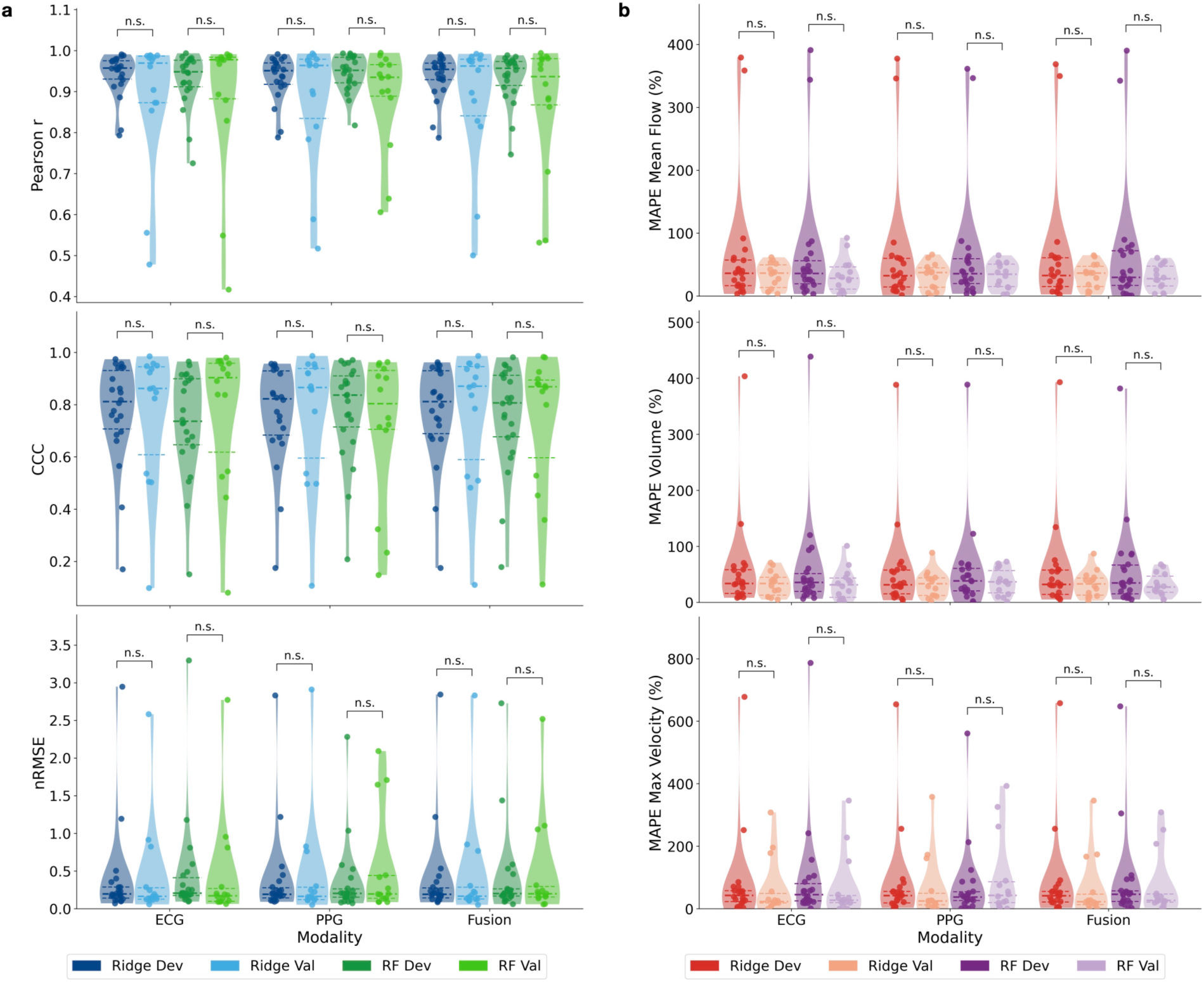
Development versus validation cohort comparison of waveform agreement and derived-metric error. Per-subject performance is shown for the development cohort (leave-one-out cross-validation, *n* = 22) and the independent validation cohort (*n* = 14) for ridge regression and random forest (RF) across three input modalities (ECG, PPG, and the multimodal fusion model [Fusion]). In each violin, the thick horizontal line denotes the median and the thinner lines denote the first and third quartiles; overlaid points show individual subjects. **a** Waveform agreement between predicted and MRI-derived ground-truth CSF flow waveforms, quantified by Pearson correlation coefficient (*r*, top), Lin’s concordance correlation coefficient (CCC, middle), and normalized root-mean-square error (nRMSE, bottom). **b** Mean absolute percentage error (MAPE) of physiological metrics derived from the predicted waveforms: mean flow (top), volume (middle), and maximum velocity (bottom). For both panels, median performance was largely preserved from development to validation, with increased dispersion and longer tails in the validation cohort reflecting a small number of poorly reconstructed subjects. Development versus validation distributions were compared within each model and modality using two-sided Mann–Whitney *U* tests; no comparison reached statistical significance (n.s.), consistent with preserved generalization, though statistical power is limited by the modest cohort sizes.

Several validation metrics equaled or modestly exceeded their development counterparts, most visibly for peak-velocity MAPE, which was lower in validation than development (for example, 22.9% versus 41.7% for the ridge fusion model; Table 3). We attribute this to two factors rather than to genuinely superior validation performance. First, the development estimates are pooled over leave-one-out folds that include two subjects the models reconstruct poorly (the worst-case behavior in Fig. 2), which widens the development distribution and inflates its aggregate error. Second, MAPE and range-normalized nRMSE are sensitive to the per-subject denominator, so cohort differences in CSF flow amplitude shift percentage error independently of reconstruction quality. We therefore interpret development and validation performance as comparable.

Consistent with testing on previously unseen subjects, the validation distributions showed greater dispersion and longer tails than development, driven by a small number of poorly reconstructed subjects (upper-quartile nRMSE reaching approximately 0.29–0.44 and lower-quartile CCC falling to approximately 0.57–0.71 in the most affected cells; Table 2). Representative validation reconstructions, including these failure cases, are shown in Supplementary Fig. 2, and the development-versus-validation comparison extended to all six feature sets is provided in Supplementary Fig. 3. For clarity, the main-text comparison in Fig. 5 is restricted to ECG, PPG, and fusion as the most informative subset (a cardiac modality, the wearable-relevant modality, and the full model), with the remaining feature sets shown in Supplementary Fig. 3.

### A minimal PPG feature set supports wearable-based reconstruction

Because PPG is widely available in consumer wearables, we tested whether CSF flow could be reconstructed from a minimal, wearable-oriented PPG feature set comprising only two physiologically interpretable feature classes: interbeat-interval variability and the pulse-amplitude ratio between consecutive beats (Fig. 6a). Across both models, this two-feature set retained waveform agreement comparable to the full 80-feature PPG set (median Pearson r approximately 0.95 versus the full-feature reference of 0.951–0.952; Fig. 6b,c). In light of the metric saturation noted above, we interpret this as evidence that a compact set of cardiovascular features retains the CSF-relevant information present in the full PPG feature set, rather than as evidence that two features are uniquely sufficient; the translational case for PPG rests on this retention together with the waveform-first peak-velocity result rather than on aggregate agreement alone.

**Fig. 6.**
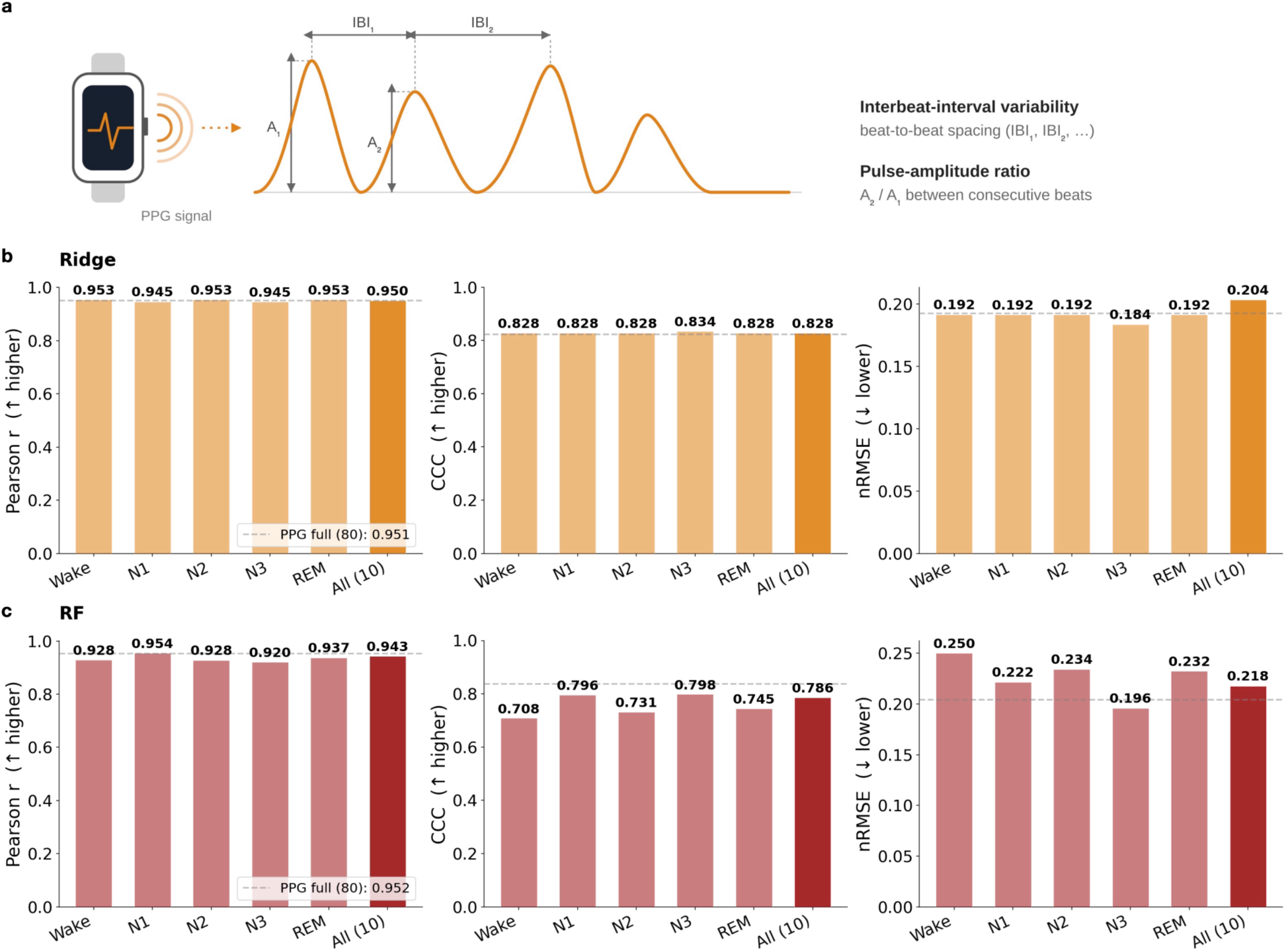
CSF flow prediction from a minimal, wearable-oriented PPG feature set. To assess the feasibility of CSF flow estimation from features obtainable on consumer photoplethysmography (PPG) devices, models were trained using only two physiologically interpretable PPG-derived feature classes: interbeat-interval variability (beat-to-beat spacing, IBI) and the pulse-amplitude ratio (the ratio of amplitudes between consecutive beats), illustrated schematically in **a**. **b, c** Per-subject waveform agreement for ridge regression (**b**) and random forest (RF, **c**), quantified by Pearson correlation coefficient (*r*, left; higher is better), Lin’s concordance correlation coefficient (CCC, middle; higher is better), and range-normalized root-mean-square error (nRMSE, right; lower is better). Within each panel, bars show performance when features were computed within individual sleep stages (Wake, N1, N2, N3, REM) and when aggregated across all stages (All), with the number of features indicated in parentheses. The dashed grey line denotes the performance of the full PPG feature set (80 features) for reference. Across both models, the minimal two-feature set retained waveform agreement comparable to the full PPG feature set, and performance was largely stable across individual sleep stages, indicating that CSF-relevant information in PPG is captured by a compact set of cardiovascular features accessible during both wakefulness and sleep.

Performance of the minimal PPG model was stable whether features were computed within individual sleep stages (Wake, N1, N2, N3, REM) or aggregated across all stages (Fig. 6b,c), indicating that the approach does not depend on capturing any particular stage and could in principle be applied to wake or partial-night recordings obtainable on consumer hardware.

### Reconstruction accuracy is largely invariant to sleep stage

We next examined whether restricting feature computation to a single sleep stage altered reconstruction accuracy across all modalities (Supplementary Fig. 4). Waveform agreement was essentially unchanged across stages for every modality and model: differences between the best-and worst-performing stages were confined to the third decimal place of Pearson r, CCC, and nRMSE (for ridge regression, per-stage median r spanned only 0.94–0.96), and the nominally best-performing stage was inconsistent across metrics and models. We therefore find no evidence of stage-specific structure in these aggregate agreement metrics; rather, CSF reconstruction is largely invariant to sleep stage. As with the modality comparison, any stage-dependent physiological structure would be expected to manifest in shape-dependent derived metrics rather than in median waveform agreement, which is saturated by shared morphology. Practically, this reinforces the wearable analysis above: accurate reconstruction does not require capturing a specific sleep stage.

### Feature importance identifies physiologically plausible predictors

To identify which inputs drove the predictions, we ranked features by the magnitude of their learned ridge weights, summed across waveform points (Fig. 7a). Within each modality, the highest-ranked features were physiologically interpretable (Fig. 7b): interbeat (mean RR) interval and heart-rate variability for ECG, pulse-geometry features (PPG waveform length) for PPG, the mean and variability of respiration rate for respiration, and interhemispheric high-gamma asymmetry for EEG. Important features were distributed across multiple sleep stages rather than concentrated in any single stage, consistent with the stage-invariance described above.

**Fig. 7.**
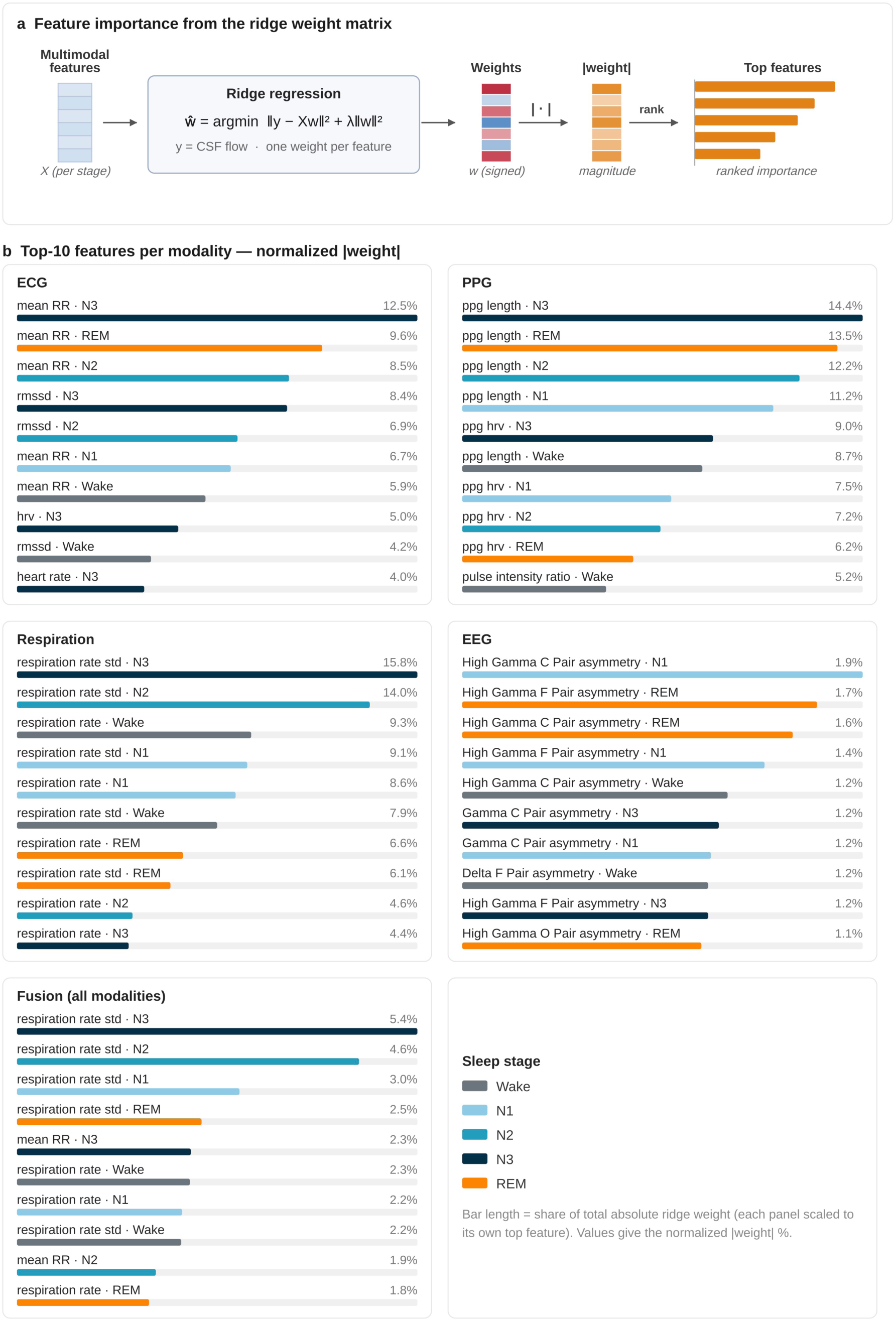
Feature importance from ridge regression weights, by modality and sleep stage. **a** Schematic of the feature-importance procedure. For each ridge regression model, the learned weight matrix (one weight per feature per CSF flow curve point) was reduced to a per-feature importance score by taking the absolute value of the weights and summing across waveform points; features were then ranked by this magnitude to identify the most influential predictors. **b** Top-10 ranked features for each input modality — ECG, PPG, respiration, EEG, and the multimodal fusion model (all modalities) — with bar length showing each feature’s share of the total absolute ridge weight, scaled within modality so each set sums to its own total, and the value at each bar giving the normalized |weight| as a percentage. Each feature reflects a specific physiological metric computed within a specific sleep stage (Wake, N1, N2, N3, REM), indicated by bar color. Across modalities, the highest-ranked features included interbeat-interval measures (mean RR interval) and heart-rate variability for ECG, pulse-geometry features (PPG waveform length) for PPG, respiration-rate mean and variability for respiration, and interhemispheric high-gamma asymmetry for EEG, with contributions distributed across multiple sleep stages rather than concentrated in any single stage.

The prominence of cardiac (RR interval, heart-rate variability) and respiratory features is consistent with the established roles of arterial pulsatility and respiration as drivers of CSF motion, lending physiological plausibility to the learned models. These rankings should be interpreted as hypothesis-generating rather than definitive: under the high-dimensional, p ≫ n regime of this study, correlated features share weight and weight-magnitude importance can vary across cross-validation folds, and the individual EEG asymmetry features each contributed only a small fraction (approximately 1–2%) of the total weight.

## Discussion

In this work we demonstrate that the temporal dynamics of CSF flow in the cerebral aqueduct can be reconstructed from noninvasive physiological signals acquired during sleep, and that this reconstruction generalizes to an independent validation cohort processed entirely by a third party. Three findings are central. First, regularized regressors recovered the cardiac-cycle CSF waveform with high fidelity across every input modality, including a minimal PPG feature set of the kind obtainable from consumer wearables. Second, deriving physiological metrics from the reconstructed waveform yielded more accurate peak-velocity estimates than predicting those metrics directly, indicating that preserving waveform morphology aids the recovery of shape-dependent quantities. Third, aggregate waveform agreement was largely saturated by the stereotyped morphology shared across healthy subjects, such that a cardiac-only baseline matched the full multimodal model on correlation-based metrics, and the physiologically meaningful, subject-specific signal was instead concentrated in the derived metrics.

The features that contributed most to prediction were physiologically interpretable and consistent with the established drivers of CSF motion. Cardiac measures (interbeat interval and heart-rate variability) and the strong performance of the heart-rate-only baseline are consistent with arterial pulsatility acting as a principal driver of CSF flow through perivascular spaces and the ventricular^4^. Respiration-rate features likewise ranked highly, in line with evidence that inspiration is a major regulator of human CSF flow^10^. That CSF dynamics can be inferred from signals recorded during sleep is further consistent with the coupling of neural slow waves, hemodynamics, and CSF oscillations observed during non-REM^11^, and with the broader role of sleep in facilitating fluid exchange and metabolite clearance^12^. The convergence of our data-driven feature rankings on these mechanisms lends physiological plausibility to the learned models.

The saturation of correlation-based agreement carries an important methodological implication: in a cohort of healthy adults whose CSF waveforms share a common systolic-peak and diastolic-trough shape, high Pearson correlation is a largely inevitable consequence of that shared morphology, and modality or sleep-stage differences cannot be resolved at the level of median waveform agreement. The clinically relevant information instead resides in shape-dependent derived quantities, such as peak velocity and stroke volume, which is precisely where waveform-first prediction conferred an advantage. This matters because aqueductal CSF stroke volume and peak velocity are parameters already used clinically to characterize altered CSF hydrodynamics, for example in the selection and monitoring of patients with idiopathic normal-pressure hydrocephalus for shunting^16^, although the predictive value of these thresholds remains debated. Our results therefore suggest that any noninvasive proxy for CSF dynamics should be evaluated on these derived metrics rather than on waveform correlation alone.

Several aspects of the analysis were designed to guard against the failure modes common to high-dimensional clinical machine learning, in which models with many more features than subjects overfit and fail to generalize. We evaluated performance on an independent validation cohort that was collected later and processed by a third party and withheld from all modelling and hyperparameter decisions. We reported concordance and error metrics (Lin’s concordance correlation coefficient and range-normalized root-mean-square error) alongside correlation, which exposed the residual amplitude error that correlation conceals. Finally, permutation testing against label-shuffled null models confirmed a statistically significant subject-specific component despite the modest sample size, providing reassurance that the predictions reflect more than the shared waveform shape.

This study has several limitations. The cohorts were small and comprised exclusively young, healthy adults, so the dynamic range of CSF flow was narrow and the models have not been tested in the disease states where noninvasive monitoring would be most valuable. The MRI and sleep-study acquisitions were performed on separate occasions rather than simultaneously, so the physiological state during imaging and during polysomnography may have differed. Because aggregate agreement metrics are saturated by shared morphology, the present data cannot establish cross-subject discrimination of CSF dynamics, and the apparent equivalence of modalities and sleep stages on these metrics should not be over-interpreted. Feature-importance estimates derived from ridge weight magnitudes are sensitive to feature correlation and to the high-dimensional regime and should be regarded as hypothesis-generating. Finally, we predicted flow only in the cerebral aqueduct; extension to the perivascular and glymphatic compartments, where much of the clearance-relevant transport occurs, remains untested.

Despite these limitations, the approach has clear potential utility. A noninvasive, low-cost, and potentially wearable estimate of CSF dynamics could enable longitudinal and at-home monitoring that is impractical with MRI, support screening and risk stratification at population scale, and make CSF physiology accessible as a repeatable measurement rather than a sparse imaging snapshot. Several next steps follow directly from the present findings. The most important is evaluation in clinical populations (idiopathic normal-pressure hydrocephalus, other hydrocephalus syndromes, and neurodegenerative diseases) to test whether predicted stroke volume and peak velocity track disease status and treatment response, building on the established clinical use of these aqueductal metrics^16^. A second priority is validation on true consumer-grade hardware, acquiring PPG and electrocardiography from wearable sensors during overnight at-home recordings, to confirm that the retention observed with the minimal PPG feature set survives realistic noise and sampling constraints. Larger and more demographically diverse cohorts, ideally with synchronous MRI and physiological acquisition, would broaden the dynamic range of CSF flow, help overcome the metric saturation observed here, and enable genuine cross-subject discrimination. Methodologically, incorporating calibrated per-subject uncertainty (for example via conformal prediction) would be valuable before any clinical use, and extending the framework to perivascular and whole-brain CSF compartments would connect this work to the glymphatic clearance pathways implicated in neurodegeneration^12^. Together, these steps would determine whether noninvasive sleep physiology can mature from the proof-of-concept demonstrated here into a validated digital biomarker of brain fluid dynamics.

## Methods

### Participants

This prospective study was approved by the Institutional Review Board (PRO00033905), and informed consent was obtained from all participants (Table 1). Data were obtained in two batches. The first batch of data was used to develop the data processing pipeline and machine learning model development. We refer to this batch of data as the development cohort, and it included 22 participants. The second batch of data was used as a validation dataset to test the model’s ability to generalize to an unseen dataset. We refer to this batch of data as the validation cohort, and it included 14 participants. All validation-cohort data were processed independently by a third party and withheld from every modelling and hyperparameter decision. Data were collected from May 2022 to May 2023. This study was structured according to the Standards for Reporting of Diagnostic Accuracy Studies (STARD)^17^, and all data are previously unpublished.

### MRI acquisition and CSF quantification

High signal-to-noise ratio, good spatial resolution, and low partial volume effects were achieved using an actively shielded 7T MRI (MAGNETOM Terra, Siemens Healthcare) with a dual-channel transmit/32-channel receive head coil (NOVA Medical). For velocity quantification, 2D phase-contrast MRI data were acquired using an in-plane resolution of 0.6 × 0.6 mm^2^, and a section thickness of 3 mm. The phase-contrast MRI measurements were performed in the cerebral aqueduct using a standard encoding velocity (V_enc_) of 15 cm/s. Retrospective cardiac triggering was provided by photoplethysmography (PPG), and the number of heart phases was kept constant at 300. Twenty images were acquired at equal intervals between the peaks of the PPG waveform. Other sequence parameters: TR/TE 105/6.38 ms, FOV 160×160 mm^2^, bandwidth 297.2 Hz/pixel, flip angle 15°, and scan time 4:09 min.

The procedure used to quantify CSF velocity in the cerebral aqueduct closely follows previous instances in the literature^18,19^. However, these methods manually outline the aqueduct and do not outline every frame of acquisition. This process assumes an unchanging aqueduct area, which has recently been shown to be false and can lead to a less accurate measurement^20^. Thus, a custom processing script was developed to automatically detect the cerebral aqueduct in all acquired frames and calculate velocity. This methodology is extensively detailed here^21^.The resulting velocity and volumetric-flow waveform, sampled at 20 points across the cardiac cycle, served as the ground-truth prediction target.

### Sleep study

Subjects completed the sleep study at the Baylor St. Luke’s Sleep Center. The Nihon Kohden Polysmith software and hardware were used to perform the sleep study. This study included recordings of the following parameters: monitoring of brain wave activity via the EEG (F3, F4, C3, C4, O1, O2) positioned as per the international 10-20 System, monitoring of blood oxygenation via the PPG, monitoring of the heart’s electrical activity via the ECG, monitoring of oronasal air flow during breathing via an air flow thermistor, and lastly the monitoring of body position throughout the recording. Each epoch was labeled with a sleep stage by a sleep expert at the Baylor College of Medicine in accordance with the American Academy of Sleep Medicine Manual for the Scoring of Sleep and Associated Events^22^. The sleep study was conducted on a separate night from the MRI acquisition.

### Data preprocessing

The signals were obtained at a sampling frequency of 200 Hz. The ECG, PPG and respiration signals were low pass filtered with a 6th order Butterworth filter with a cutoff frequency of 50 Hz, which eliminates powerline interference, electromyographic noise, and electrode motion artifact noise^23^. Normal filtering is done only in the forward direction in time. This causes a time delay and thus a change in the phase of the signal. We can easily fix that by also filtering the signal backwards in time^24^. This is called a zero-phase filter and is beneficial here since we want to preserve all information in time without introducing distortions. The EEG signal was high-pass filtered, with the same filter mentioned prior, at a cutoff frequency of 0.3 Hz to eliminate electrode drift. The EEG was also notch filtered at 60 Hz to eliminate powerline noise (stop band width of 2 Hz around 60 Hz). We did not have an upper cutoff frequency so that we could analyze the gamma and high gamma band of the EEG. Note that we can only resolve frequencies up to 100 Hz due to the Nyquist theorem^25^. Next, we segment the data into 30 s non-overlapping blocks aligned with the sleep scoring labels. All segmented blocks associated with the same sleep stage are grouped together for feature extraction. This allowed us to analyze how the performance of our regressor changes with different sleep stages. From the segmented and grouped blocks of data we performed feature extraction. For each modality we additionally computed a common set of nonlinear complexity features (signal entropies, fractal dimensions, Hjorth parameters, and the Hurst exponent)^26–29^. In addition, modality specific features were calculated.

The ECG is a non-invasive tool for assessing the health of the heart. In this work, we only use one lead of the ECG signal. We calculate the heart rate, mean R wave peak to peak distance (RR interval), heart rate variability, root mean square of successive differences between normal heartbeats, percentage of successive RR intervals that differ by more than 50ms, low frequency and high frequency power, the ratio of low frequency (LF) to high frequency (HF) power, and the normalized low and high frequency power^30^. These features are calculated for every epoch and are then aggregated and averaged over the five sleep stages. This strategy is implemented for all four physiological modalities. The total number of features for the ECG feature set was 110.

PPG is a simple optical technique that detects fluctuations in blood volume by measuring reflected red or infra-red light. This reflected light is proportional to changes in blood volume via the Beer-Lambert law^31^. For the PPG we calculated the pulse intensity ratio, heart rate variability, percent of dicrotic waves, and the length of the PPG waveform (arc length of PPG curve per beat then averaged across epoch). The total number of features for the PPG feature set was 80.

The respiration signal detects the breathing pattern of the subject over time. From this waveform we calculated the mean and standard deviation of the respiration rate, the maximum inhalation slope, maximum exhalation slope, and the area under the respiration curve. The total number of features for the respiration feature set was 85.

The EEG is a powerful tool for studying the electrical behavior of the brain^32^. This modality is non-invasive and relatively easy to obtain as open-source platforms and consumer devices for the measurement of EEG continue to proliferate^33^. From the EEG we calculate the power in six frequency bands (delta, theta, alpha, beta, gamma, and high gamma) per EEG channel, ratios between the power in the six frequency bands across channels, asymmetry metrics between homologous pairs of EEG electrodes, correlation graph metrics, mutual information graph metrics, and coherence graph metrics. Due to the numerous channels and the higher complexity of feature extraction, we end up with 2,170 features for the EEG dataset.

As a physiologically naive baseline, we additionally defined a minimal feature set consisting only of summary heart-rate and respiration-rate statistics (10 features in total), aggregated across the five sleep stages. Because cardiac pulsatility is itself a driver of CSF motion, this baseline quantifies the performance attainable from heart rate alone and serves as a reference against which the richer feature sets are judged. Finally, the four physiological feature sets were concatenated to form a full-modality fusion feature set (2,445 features). For every feature set the number of features p exceeded the number of participants n, situating the problem in the high-dimensional p >> n regime and motivating the regularized and ensemble models described below^34,35^.

### Predictive modelling framework

We framed CSF flow estimation as a supervised regression problem. For each input feature set, the model mapped the per-participant feature vector to the 20-point CSF flow waveform measured by PC-MRI, so that the target matrix Y had dimension n x 20 (n participants by 20 cardiac-cycle points) and the feature matrix X had dimension n x p. We trained two complementary regressors per feature set: ridge regression, a linear model with an L2 penalty on the coefficients that is well suited to the p >> n setting^36^, and random forest, a nonlinear ensemble of regression trees^37^. Each model has a small number of hyperparameters (the ridge regularization strength, and the random-forest tree count and depth), which were tuned by grid search using only the development cohort. Models were implemented in Python with scikit-learn ^38^.

We considered two prediction strategies. In the waveform-first strategy (Mode 1), the model predicted the full 20-point waveform and the physiological metrics of interest were then computed from the reconstructed waveform. In the direct strategy (Mode 2), separate models were trained to predict each scalar metric directly from the same features. Comparing the two strategies isolates the benefit of preserving waveform morphology when estimating derived metrics.

Two evaluation regimes were used. Within the development cohort, performance was estimated by leave-one-out cross-validation: the model was trained on all but one participant and tested on the held-out participant, repeated for every participant. To assess generalization, the models, preprocessing, and hyperparameters were then fixed on the development cohort and applied without modification to the independent validation cohort. As a reference, we computed a mean-prediction baseline in which the average of the training waveforms was used as the prediction for every test participant.

### Evaluation metrics

Waveform reconstruction was assessed per participant using three complementary metrics computed between the predicted and ground-truth waveforms. The Pearson correlation coefficient (r) quantifies linear association but is insensitive to differences in amplitude and offset^39^. Lin’s concordance correlation coefficient (CCC) additionally penalizes departures from the line of identity, capturing amplitude and scale error^40^. The range-normalized root-mean-square error (nRMSE) was defined as the root-mean-square error between the predicted and ground-truth waveforms divided by the peak-to-trough amplitude (maximum minus minimum across the 20 points) of each participant’s ground-truth waveform, such that a value of 0.20 corresponds to an error equal to 20% of that participant’s CSF flow amplitude.

From each waveform we derived three physiologically meaningful metrics: mean flow (the average flow over the cardiac cycle), volume (the integrated flow over the cycle), and peak velocity (the maximum velocity). Agreement between metrics derived from the predicted and ground-truth waveforms was quantified by the mean absolute percentage error (MAPE). Unless otherwise stated, all metrics are reported as the median and interquartile range across participants.

### Statistical analysis

To test whether reconstruction accuracy exceeded chance, we performed permutation (null-model) testing: participant labels were randomly shuffled and the models retrained, repeating this procedure 100 times to build a null distribution of the mean Pearson r, against which the observed value was compared to obtain an empirical p-value. To compare the waveform-first and direct prediction strategies, we computed the paired per-participant difference in MAPE (delta-MAPE) and estimated 95% confidence intervals by bootstrap resampling, treating a difference as significant when its interval excluded zero. Development and validation distributions were compared for each model and modality using two-sided Mann-Whitney U tests, which assume neither paired samples nor normality. Statistical significance was defined as p < 0.05.

### Individual sleep stage prediction

Because features were computed separately within each sleep stage, we additionally trained and evaluated models using features from a single stage at a time (Wake, N1, N2, N3, or REM) for the ECG, PPG, respiration, EEG, naive-HR, and fusion feature sets, using ridge regression and random forest under the same leave-one-out scheme. This analysis quantified the dependence of reconstruction accuracy on the sleep stage from which features were derived (Supplementary Fig. 4).

### Wearable PPG simulation

To assess feasibility on consumer-grade hardware, we constructed a minimal PPG feature set restricted to two physiologically interpretable, wearable-obtainable feature classes: interbeat-interval variability and the pulse-amplitude ratio between consecutive beats. Models were trained on this minimal set using features computed within each individual sleep stage and aggregated across all stages, and performance was compared against that of the full 80-feature PPG set.

### Feature importance

To identify the most influential predictors, we examined the learned ridge weight matrix (dimension p x 20). For each feature we took the absolute value of its weights and summed across the 20 waveform points to obtain a single importance score, yielding a per-feature importance vector of length p^41,42^. Within each modality, features were ranked by this score and the ten highest-ranked features retained; in the corresponding figure, bar length expresses each feature’s share of the total absolute weight within its modality. Because weight-magnitude importance can be sensitive to feature correlation in the high-dimensional regime, these rankings are interpreted as descriptive rather than definitive.

## Code and data availability

The code used for data processing, feature extraction, model training, and analysis is available from the corresponding author upon reasonable request. De-identified data supporting the findings of this study are available from the corresponding author upon reasonable request, subject to institutional data-sharing agreements and the governing ethical approvals.

