## Supplemental Material for "Multimodal Sleep Physiology Reconstructs Cerebrospinal Fluid Dynamics: A Candidate Digital Biomarker from Noninvasive Sensing"

### Supplemental Figures

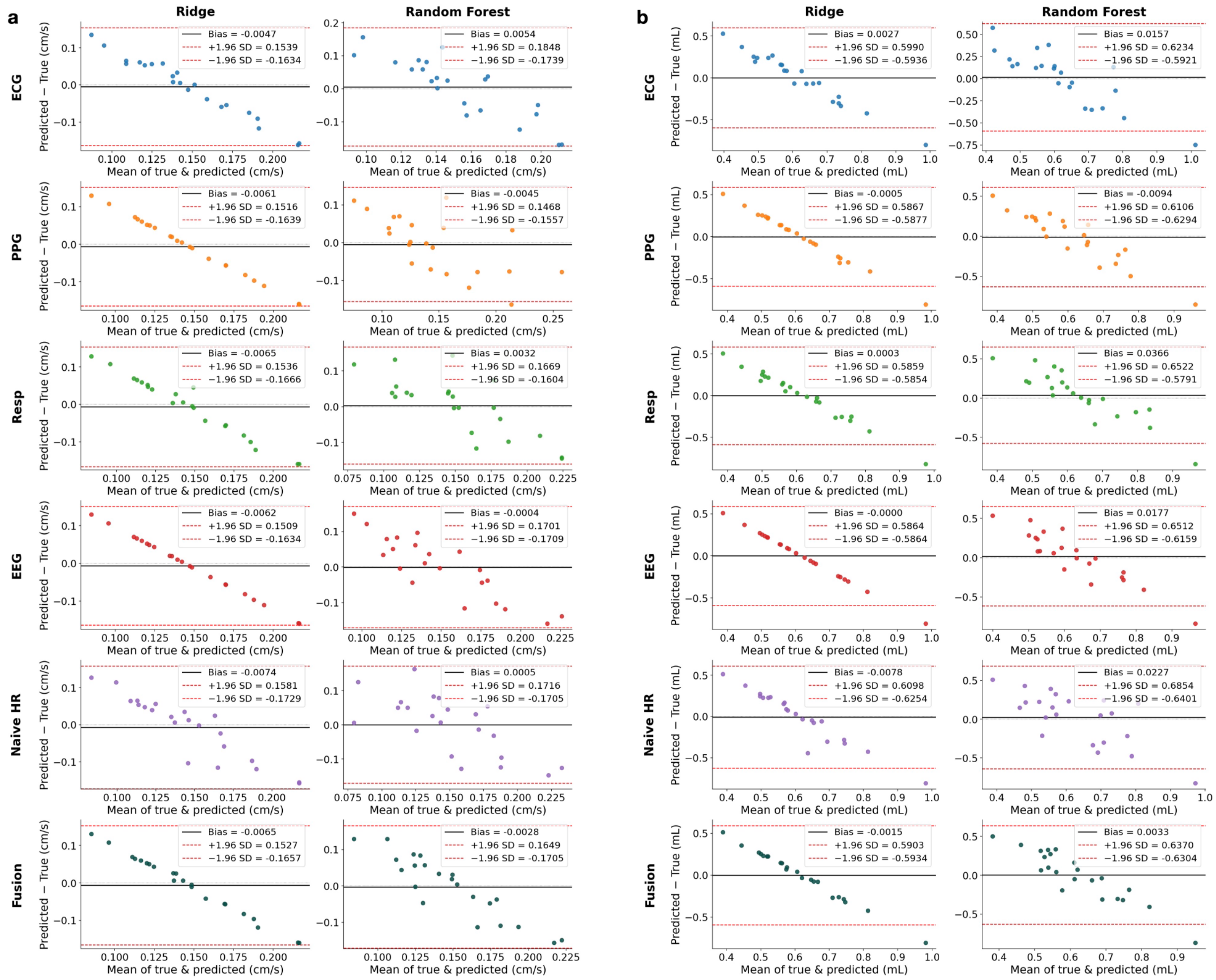


**Supplementary Fig. 1 | Bland–Altman agreement for derived volume and peak velocity.** Bland–Altman plots comparing waveform-derived CSF volume and peak (maximum) velocity against the MRI-derived ground truth for ridge regression and random forest across all input feature sets; the corresponding analysis for mean flow is shown in main-text Fig. 3a. Solid lines denote the mean bias and dashed lines the ±1.96 SD limits of agreement.

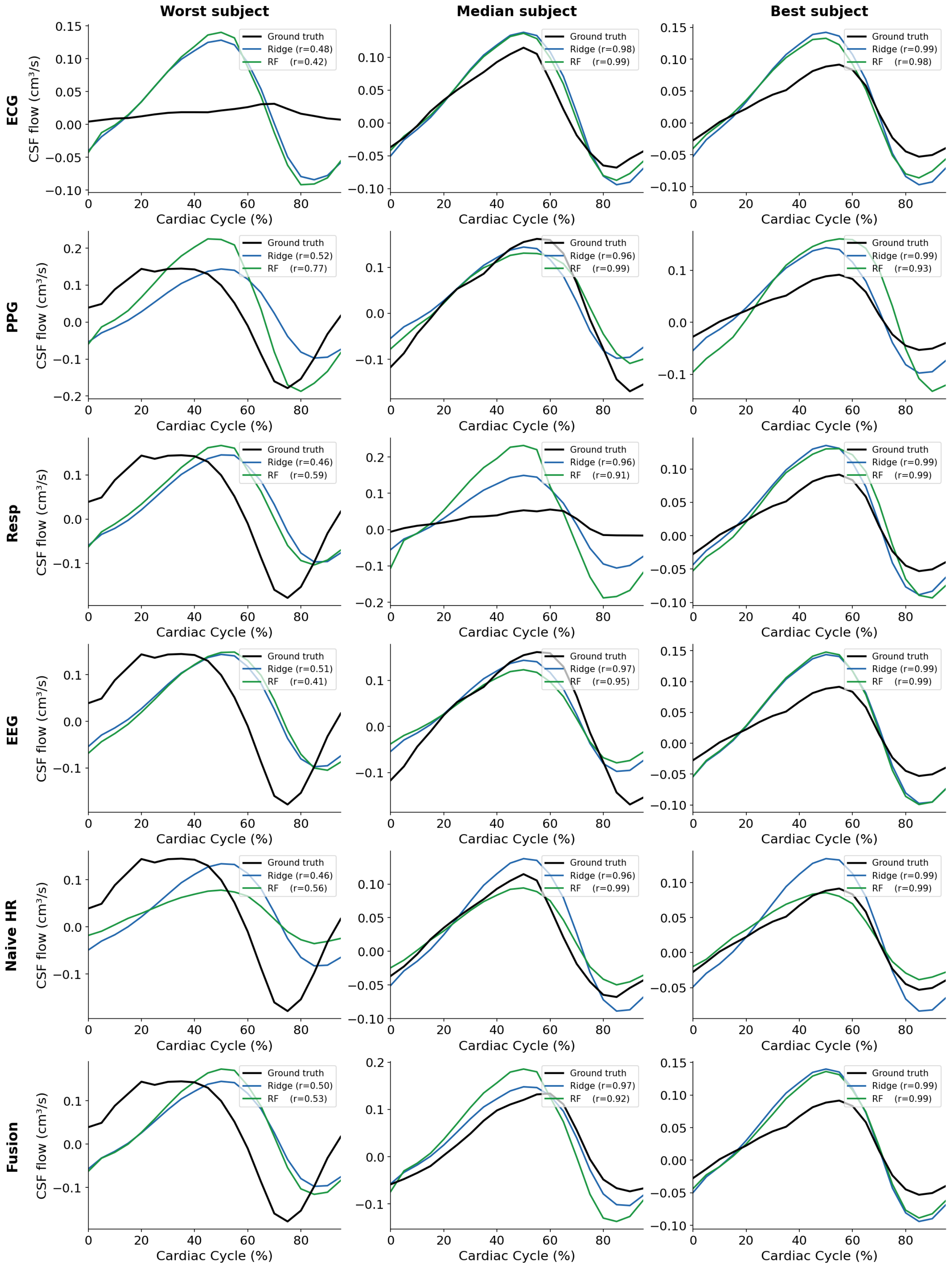
Worst subject
Median subject
Best subject
ECG
PPG
Resp
EEG
Naive HR
Fusion
CSF flow (cm³/s)
Cardiac Cycle (%)
Ground truth
Ridge (r=0.48)
RF (r=0.42)
Ground truth
Ridge (r=0.98)
RF (r=0.99)
Ground truth
Ridge (r=0.99)
RF (r=0.98)
Ground truth
Ridge (r=0.52)
RF (r=0.77)
Ground truth
Ridge (r=0.96)
RF (r=0.99)
Ground truth
Ridge (r=0.99)
RF (r=0.93)
Ground truth
Ridge (r=0.46)
RF (r=0.59)
Ground truth
Ridge (r=0.96)
RF (r=0.91)
Ground truth
Ridge (r=0.99)
RF (r=0.99)
Ground truth
Ridge (r=0.51)
RF (r=0.41)
Ground truth
Ridge (r=0.97)
RF (r=0.95)
Ground truth
Ridge (r=0.99)
RF (r=0.99)
Ground truth
Ridge (r=0.46)
RF (r=0.56)
Ground truth
Ridge (r=0.96)
RF (r=0.99)
Ground truth
Ridge (r=0.99)
RF (r=0.99)
Ground truth
Ridge (r=0.50)
RF (r=0.53)
Ground truth
Ridge (r=0.97)
RF (r=0.92)
Ground truth
Ridge (r=0.99)
RF (r=0.99)

**Supplementary Fig. 2 | Representative CSF waveform reconstructions in the validation cohort.** As in Fig. 2 but for the independent validation cohort, showing the MRI-derived ground-truth and predicted CSF flow waveforms (ridge regression and random forest) for the worst-, median-, and best-performing subjects across each input modality.

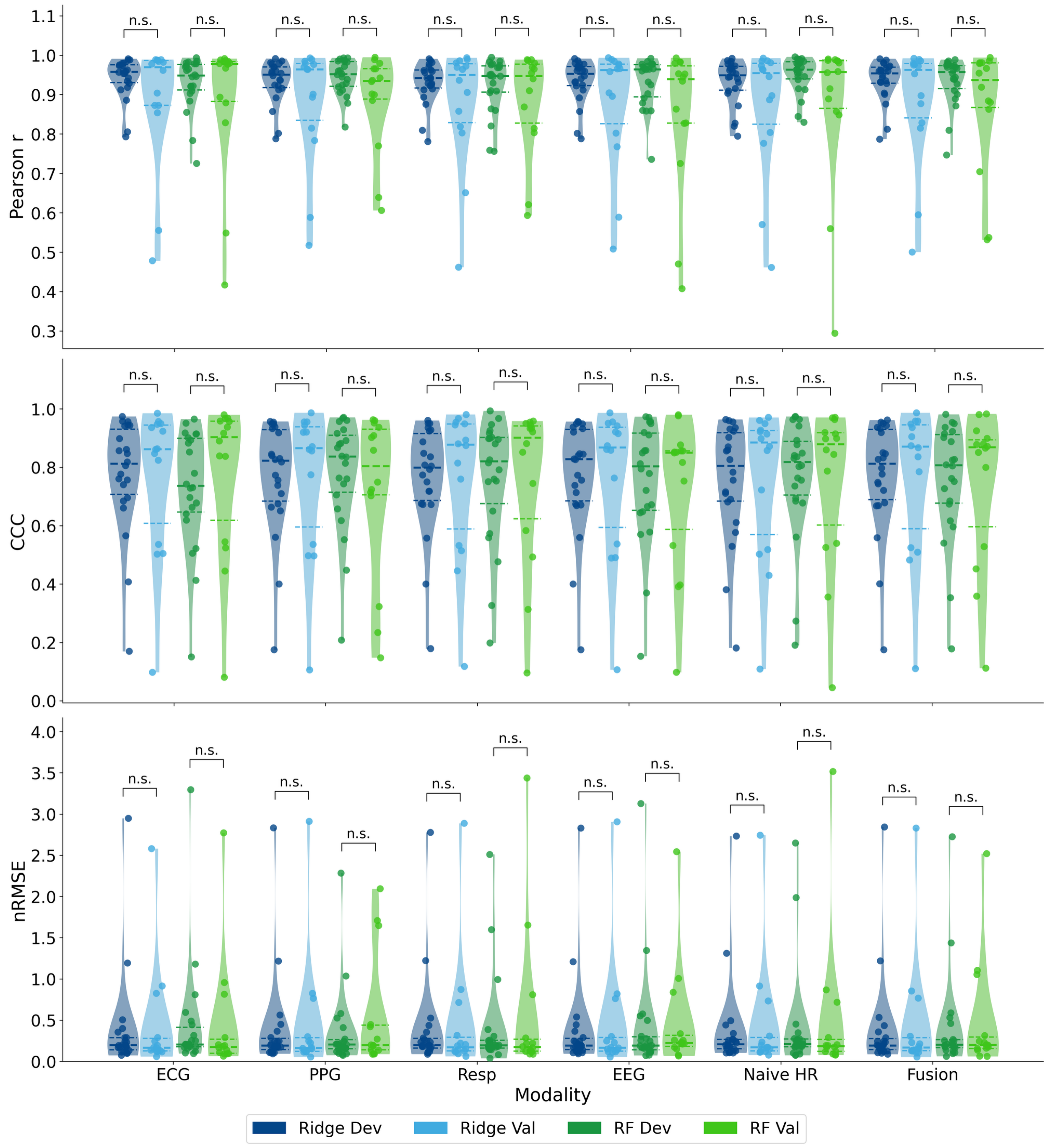


**Supplementary Fig. 3 | Development versus validation comparison across all feature sets.** Extension of Fig. 5 to all six input feature sets (ECG, PPG, respiration, EEG, the naïve heart-rate baseline, and fusion), showing per-subject Pearson r, CCC, and nRMSE for the development and validation cohorts under ridge regression and random forest.

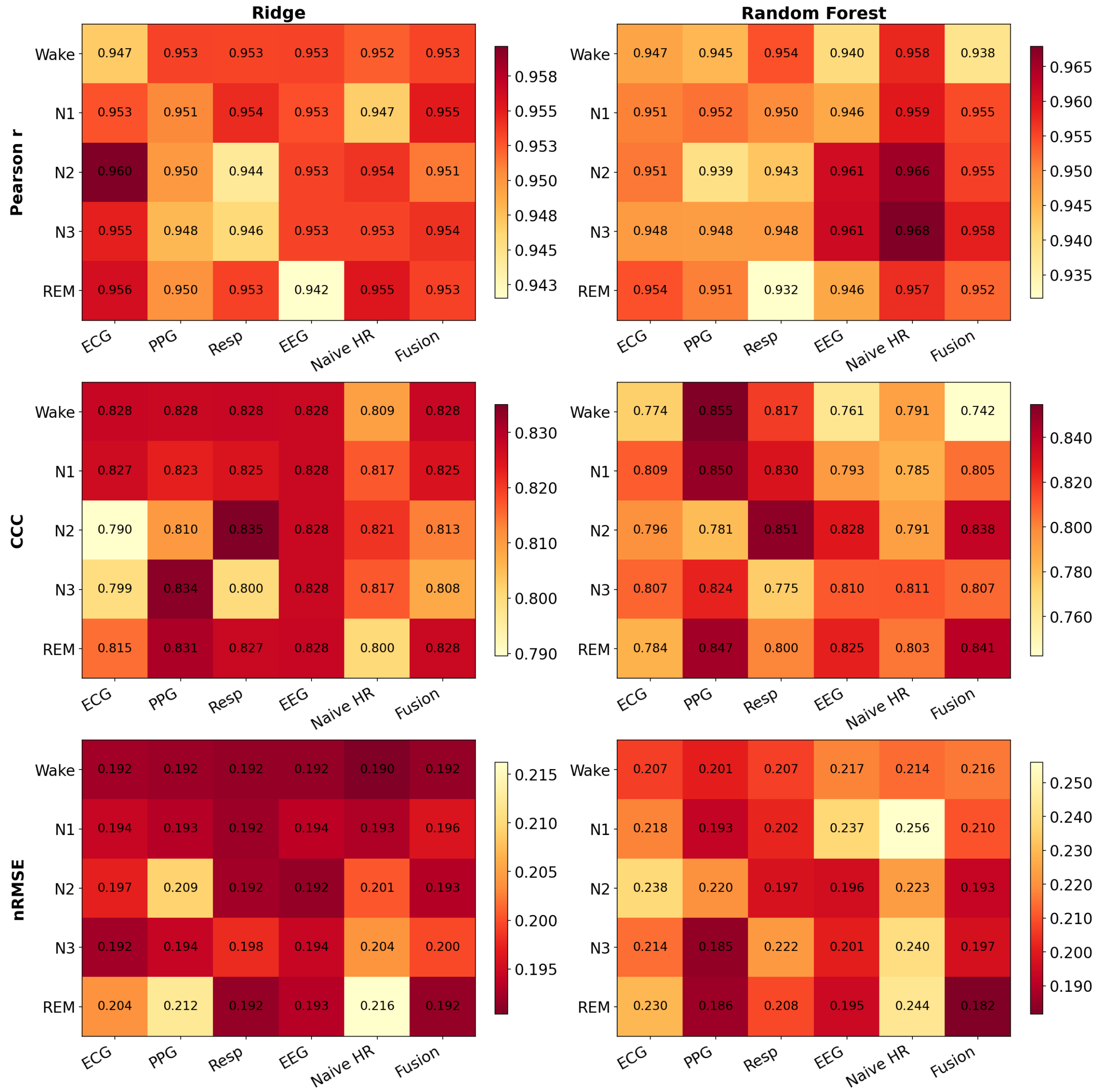


**Supplementary Fig. 4 | Reconstruction accuracy by sleep stage.** Per-stage waveform agreement (Pearson r, CCC, and nRMSE) for ridge regression and random forest across all six feature sets, with features computed within each individual sleep stage (Wake, N1, N2, N3, REM). Note the narrow per-panel color scales: differences across stages are confined to the third decimal place, indicating that reconstruction accuracy is largely invariant to sleep stage.
